# Booster and anergic effects of the Covishield vaccine among healthcare workers in South India

**DOI:** 10.1101/2021.08.04.21261601

**Authors:** Priya Kannian, Pasuvaraj Mahanathi, Veeraraghavan Ashwini, Nagalingeswaran Kumarasamy

**Affiliations:** Department of Clinical Research, The Voluntary Health Services Hospital, Chennai, Tamil Nadu, India; VHS-Infectious Diseases Medical Centre, The Voluntary Health Services Hospital, Chennai, Tamil Nadu, India

**Keywords:** Covishield, anti-SARS-CoV2 spike antibody, healthcare workers, booster, anergy

## Abstract

Covishield (same as ChAdOx1) vaccine was rolled out in January 2021 against SARS-CoV2 in India. Although studies show good efficacy after two doses, there is limited data on the fate of the elicited antibody responses over time in groups with or without prior exposure to SARS-CoV2. Therefore, in this study we proposed to test naïve or previously exposed healthcare workers (HCWs) longitudinally after both doses for anti-SARS-CoV2 spike antibody (ASSA) levels. Serum samples were collected from 205 HCWs at days 14 and 28 after first dose, and at days 14, 28 and 3-months after second dose. ASSA levels were quantitated by ECLIA method. Non-responder rate was 17% (35/205) on day 14 and 2% (5/205) on day 28 after the first dose. After the second dose, the responder rate was 100%. Non-responder rate was significantly higher among males (p<0.00001) and senior citizens (p=0.008). The second dose boosted a 27-fold increase in the COVID-19 naïve (CN) group, but caused a 1.5-fold decline in the previously exposed groups. By three months, the antibody levels declined 3-4-folds in all the groups. In spite of high antibody levels (GM-1007 U/ml) after the second dose, 14% developed mild breakthrough infections (BTI). The booster effect was significantly higher when given 10-14 weeks later. The responder rate for Covishield was 98% after first dose and 100% after second dose. The vaccine elicited a prime-boost effect in CN HCWs and a boost-anergy effect in the previously exposed HCWs. ASSA levels began to decline proportionately by three months.

## Introduction

The COVID-19 pandemic caused by SARS-CoV2 caused high morbidity and mortality in India. The COVID-19 vaccination programme was rolled out in the end of January 2021. Two different vaccines are administered in India - Covishield™ and Covaxin™. Covishield™ is a chimapanzee adenoviral vector carrying the genetic material coding for the spike protein. This is the same as the Astra Zeneca’s ChAdOx1. It is manufactured at Serum Institute, India and is the most widely administered vaccine. Covaxin™ is an inactivated whole virus vaccine manufactured by Bharath Biotech Ltd. Few studies from India have shown the effectiveness of Covishield – reduction in the incidence rate by 93% in the vaccinated (15.23 lakhs) compared to the unvaccinated (72,283) Indian Armed Forces cohorts [1]; significant reduction in transmission among 3196 institutional employees [2]; reduced mortality in the fully vaccinated (12.5%) compared to the unvaccinated cohorts (31.5%) among 1168 moderate to severe COVID-19 patients hospitalized during the second wave [3]; and reduced hospitalization in the fully vaccinated air warriors compared to the unvaccinated counterparts [4].

Understanding the antibody responses elicited by the vaccine is important to decide upon the necessity and the time gap for booster doses. A number of studies particularly from the United Kingdom have compared the antibody responses elicited by the Pfizer’s mRNA vaccine, BNT162b2 and the Astra Zeneca’s ChAdOx1 vaccine. Lumley *et al* have shown that two doses of either of these vaccines elicit antibody responses proportional to the natural infection; and the protection against infection is robust including the B.1.1.7 variant [5]. Two groups compared the antibody responses elicited by BNT162b2 and ChAdOx1 in cross-sectional studies. The sampling time after the vaccination varied widely in these two large cohort studies allowing time gap analyses of the elicited antibody responses in age-matched groups. They have concluded that ChAdOx1 elicited lower and slower antibody responses compared to BNT162b2, which declined over time [6,7]. The major limitation in these studies is that the decline seen in the antibody responses over time is not in the same individuals. There is limited follow up data on the antibody responses against ChAdOx1 unlike BNT162b2 within the same cohort, and only small groups of vaccinees have been studied after the second dose of ChAdOx1.

In the current study, we propose to determine the antibody responses at two time points after each of the two doses of Covishield among healthcare workers (HCWs), and one three-month time point after the second dose. We have analysed the rising and declining trends of the elicited antibody responses among those who were previously exposed and not exposed to SARS-CoV2 in a longitudinal fashion up to three months after the second dose.

## Materials and Methods

### Patients and samples

The study was approved by the VHS-Institutional Ethics Committee (Proposal# VHS-IEC/72-2020). A total of 50 post-COVID patients (PCPs) tested for anti-SARS-CoV2 spike antibodies (ASSA) were analysed retrospectively. HCWs (n=220) were recruited on the 14^th^ day post-first vaccine dose of Covishield (same as ChAdOx1 manufactured in India) after a written informed consent. Serum samples were collected at the time of recruitment. Demographic details, past COVID-19 exposure and vaccination dates were collected. Longitudinal serum samples were collected on 28^th^ day after the first dose, and 14^th^ day, 28^th^ day and 3^rd^ month after the second dose. The details of the number of samples tested at each of the longitudinal time points, basic inclusion criterion and the reasons for fall-outs are depicted in figure 1. Serum samples were stored at -80ºC until further use.

**Figure 1:**
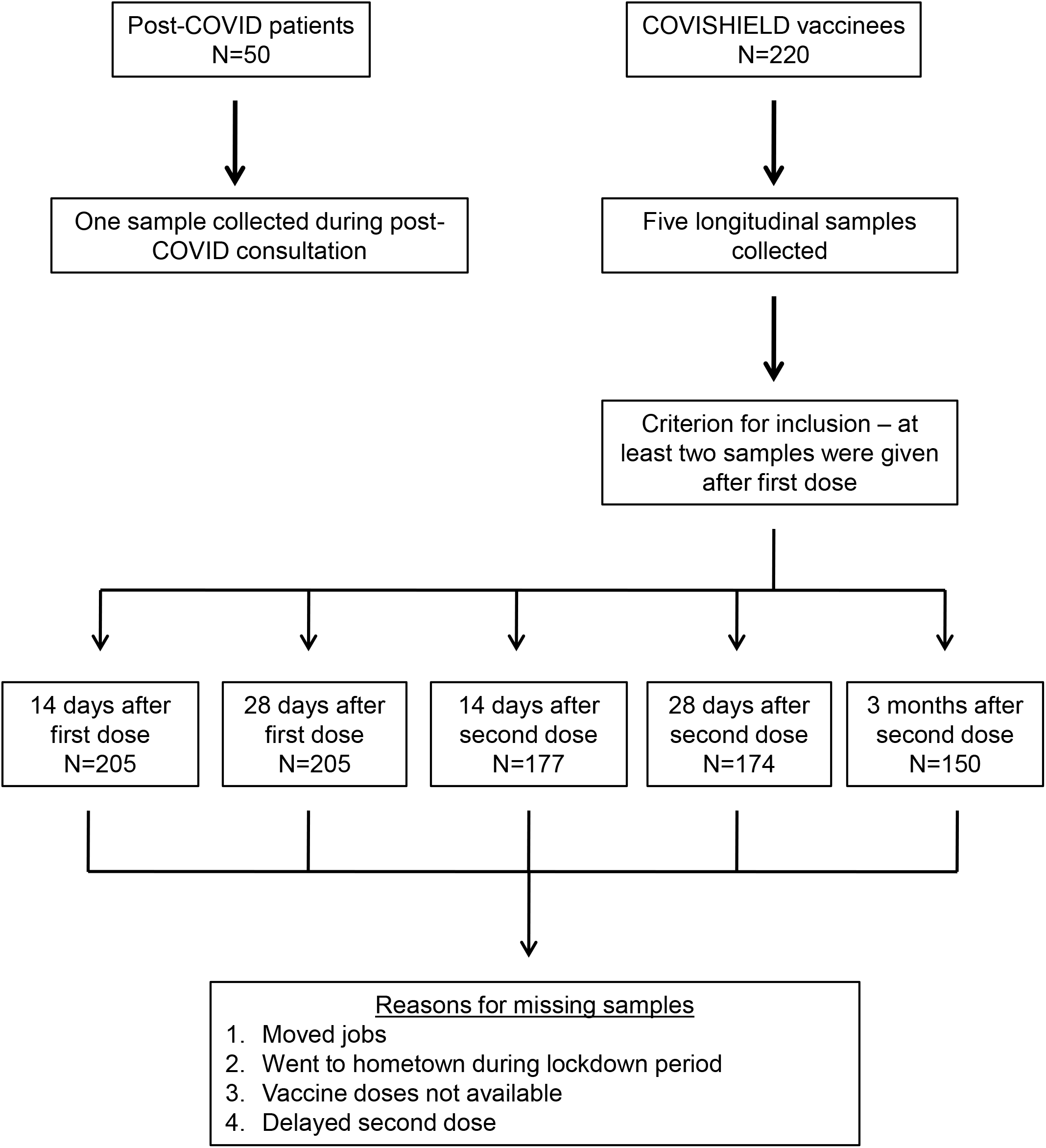
Flow chart depicting the cohorts, sample collection time points, criterion for inclusion and reasons for missing samples.

### Anti-SARS-CoV2 spike antibodies (ASSA)

Anti-SARS-CoV2 spike antibodies (ASSA) were detected by electrochemiluminescence assay (ECLIA) using Cobas e411 automated analyser (Roche, Germany). The analytical range of the reagent is 1-250 U/ml. Values less than 1 U/ml were considered negative. Samples with values > 250 U/ml were diluted to 1:400 with 1x Dulbecco’s phosphate buffered saline (PBS; HiMedia, India) and the final antibody concentration was determined.

### Statistical analysis

The mean, median and quartile values were calculated using Microsoft excel. Chi-square test and Anova tests were done using free online calculators from VassarStats and Social Science Statistics.

## Results

ASSA were detected in 83% (170/205) on the 14^th^ day and 98% (200/205) on the 28^th^ day after the first vaccine dose among the HCWs. All the PCPs had detectable antibodies (GM – 209 U/ml; range: 1.3-6424 U/ml). A small percentage of the HCWs did not develop ASSA after the first dose and were defined as non-responders (Table 1). However, all of them developed ASSA responses after the second dose. More females (n=150) participated in the study than males (n=55). Yet, the non-responder rate was higher among males than females. This difference was statistically significant (p<0.00001; Chi-square test). Age-wise analysis indicated a higher non-responder rate among the senior citizens, which was also statistically significant (p=0.008; Chi-square test).

**Table 1:** Non-responder rates after the first vaccine dose.

|  | **Total**<br>**N (%)** | **14 days**<br>**n (%)** | **28 days**<br>**n (%)** | **p value**<br>**(Chi-square test)** |
| --- | --- | --- | --- | --- |
| Overall | 205 | 35 (17) | 5 (2) |  |
| <u>Gender</u> |  |  |  |  |
| Males | 55 (27) | 14 (25) | 3 (5) | < 0.00001 |
| Females | 150 (73) | 21 (14) | 2 (1) |  |
| <u>Age (years)</u> |  |  |  |  |
| 20-44 | 141 (69) | 13 (9) | 2 (1) | 0.008 |
| 45-60 | 51 (25) | 15 (29) | 1 (2) |  |
| >61 | 13 (6) | 7 (54) | 2 (15) |  |

N – total number of participants in each group.
n – number of non-responders.
% - percentage of non-responders over the total number of participants.

The 205 HCWs were stratified into four groups – COVID-19 naïve (CN; did not develop any symptoms of COVID-19), mild COVID prior (MCP; confirmed RT-PCR-positive HCWs in 2020), probable exposure prior (PEP; did not develop any symptoms of COVID-19, but the ASSA levels were as high as the MCP group on the 14^th^ day after the first vaccine dose suggesting asymptomatic exposure), and breakthrough infections (BTI; developed COVID-19 symptoms either after the first or second dose of vaccination and were tested RT-PCR positive). The gender-wise (p=0.08; Chi-square test) and age-wise (p=0.5; Chi-square test) analyses of the ASSA responses between these four groups did not show any statistically significant differences (Table 2).

**Table 2:** Anti-SARS-CoV2 spike antibodies elicited after the first and second doses of Covishield vaccine in the four groups of healthcare workers.

|  | **Total** | **COVID**<br>**naïve** | **Probable**<br>**exposure**<br>**prior** | **Mild COVID**<br>**prior** | **Break**<br>**through**<br>**infections** |
| --- | --- | --- | --- | --- | --- |
| Overall n (%) | 205 | 88 (43) | 76 (37) | 13 (6) | 28 (14) |
| <u>Gender - n (%)</u> |  |  |  |  |  |
| Males | 55 | 27 (31) | 13 (17) | 4 (31) | 11 (39) |
| Females | 150 | 61 (69) | 63 (83) | 9 (69) | 17 (61) |
| <u>Age in years – n (%)</u> |  |  |  |  |  |
| 20-44 | 141 | 57 (65) | 54 (71) | 12 (92) | 18 (64) |
| 45-60 | 51 | 23 (26) | 20 (26) | 0 | 8 (29) |
| >61 | 13 | 8 (9) | 2 (3) | 1 (8) | 2 (7) |
| <u>After first dose</u> |  |  |  |  |  |
| 14 days | 205 | 12 | 16549 | 20407 | 9 |
| GM (range) |  | (1-244) | (920-68360) | (4732-88400) | (1-440) |
| 28 days | 205 | 61 | 10766 | 14261 | 68 |
| GM (range) |  | (2-1012) | (856-39652) | (3724-60880) | (1.7-608) |
| <u>After second dose</u> |  |  |  |  |  |
| 14 days | 177 | 1206 | 9728 | 13584 | 1149 |
| GM (range) |  | (47-16084) | (1980-77160) | (2692-64920) | (138-11888) |
| 28 days | 174 | 870 | 7977 | 12039 | 1007 |
| GM (range) |  | (29-12824) | (1720-34908) | (3032-37476) | (90-27848) |
| 3 months | 150 | 306 | 4543 | 6545 | 14840 |
| GM (range) |  | (16-2660) | (1124-37200) | (1376-22004) | (1744-89880) |

### Four-fold decline in ASSA levels

Among the 88 CN HCWs, 62 (71%) elicited ASSA responses by the 14^th^ day (GM – 12 U/ml; Table 2) and 83 (94%) elicited ASSA responses by the 28^th^ day (GM – 61 U/ml) after the first prime dose. All the 88 HCWs had elicited detectable antibody responses after the second dose. Similarly among the 28 HCWs who developed BTI after the vaccination, 19 (68%) developed antibodies by the 14^th^ day (GM – 9 U/ml) after the first dose, while all the 28 HCWs developed antibody responses by the 28^th^ day (GM – 68 U/ml). The second vaccine dose elicited a 27-fold and 17-fold increase in ASSA levels by the 14^th^ day in the CN (GM – 1206 U/ml) and BTI (GM – 1149 U/ml) groups, respectively. By the 28^th^ day, the ASSA levels began to decline in both the groups, which are depicted as 27-75% interquartile ranges (IQR) in figures 2A and 2D. However, at three months post-second dose there is a statistically significant difference (p<0.00001; Table 3) in the ASSA levels between these two groups. HCWs in the CN group showed a four-fold decline at three months, while those in the BTI group showed a 15-fold increase due to the development of COVID-19. A similar 3-4-fold decline was seen in the ASSA levels in the MCP and PEP groups at three months (Table 2).

**Table 3:**
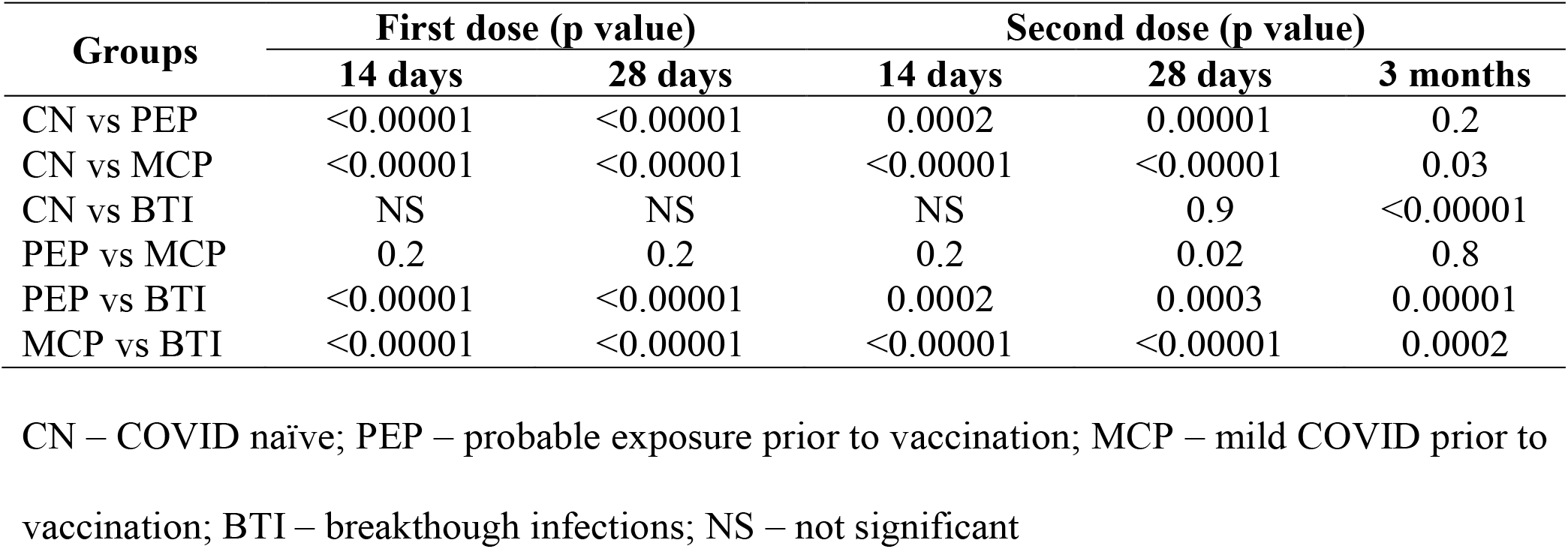
Difference in anti-SARS-CoV2 spike antibody levels between the four groups of healthcare workers by Tukey’s Anova test.

**Figure 2:**
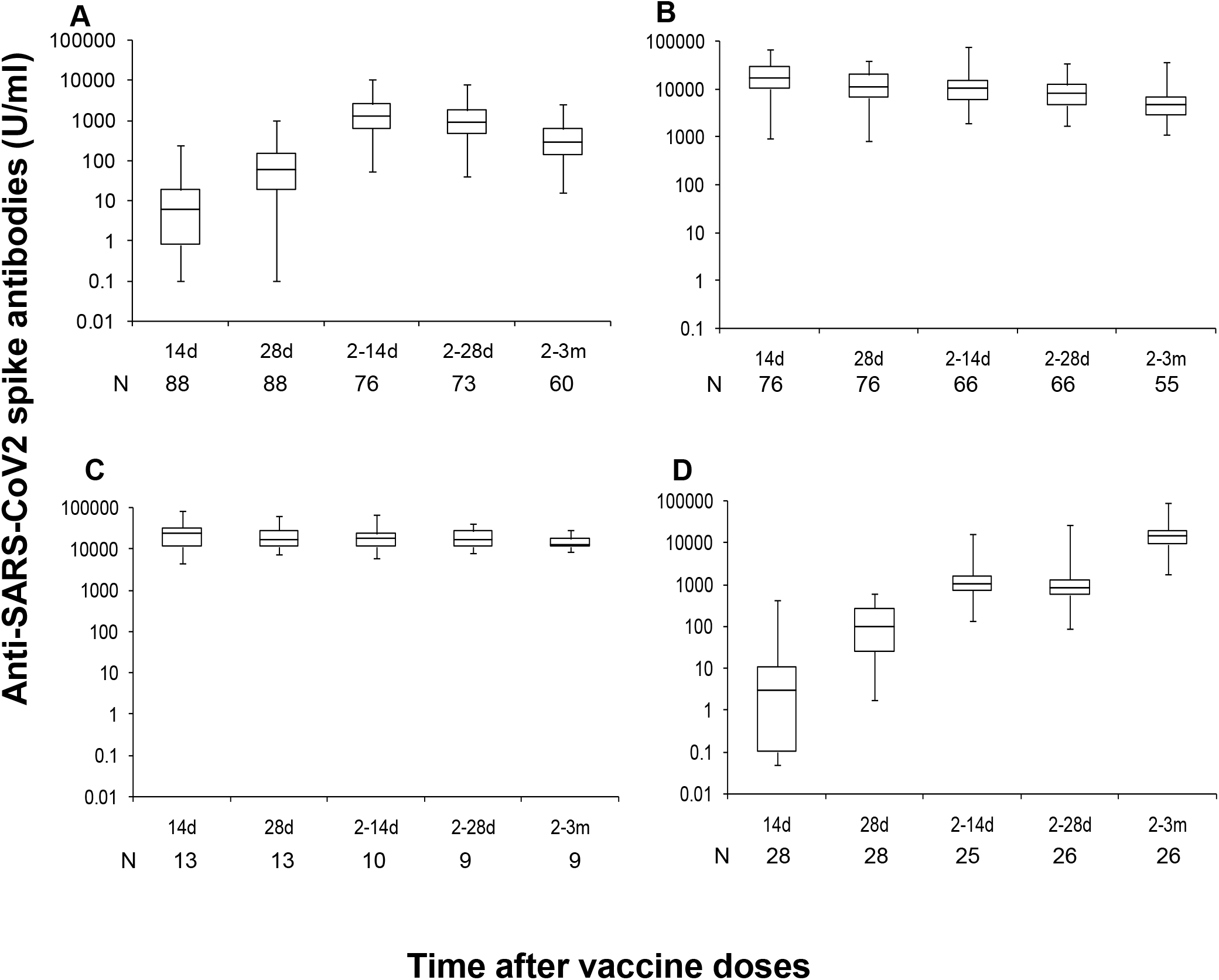
The 25-75% interquartile range (IQR) trends for antibody responses elicited over time in the four groups of vaccinees. The X-axis denotes the sample tested time points – 14d: 14 days after first dose; 28d: 28 days after first dose; 2-14d: 14 days after second dose; 2-28d: 28 days after second dose; 2-3m: three months after second dose. N is the number of samples tested at each time point. The Y-axis denotes the anti-SARS-CoV2 spike antibody concentration in logarithmic U/ml. **A**. COVID-19 naïve group (CN). **B**. Probable exposure prior (PEP) group. **C**. Mild COVID-19 prior (MCP) group. **D**. Breakthrough infections (BTI) group.

### Vaccine boosted the memory and caused anergy in the MCP and PEP groups

ASSA levels were very high among the HCWs in the MCP (GM – 20407 U/ml) and PEP (GM – 16549 U/ml) groups after the 14^th^ day of the first vaccine dose (Table 2). Their ASSA levels remained consistently high throughout the longitudinal study period (Figures 2B and 2C). The ASSA levels were significantly higher compared to the CN and BTI groups (Table 3). Although the 25-75% IQR trends were the same in the MCP and PEP groups (Figures 2B and 2C), they were significantly different from the CN and BTI groups (Figures 2A and 2D). These differences were statistically significant (Table 3). Although not statistically significant, the ASSA levels in the PEP group were slightly lower than the MCP group at most of the time points. This difference was statistically significant on the 28^th^ day after the vaccine dose (Table 3). Additionally, the GM ASSA levels clearly indicate that the first dose boosted the memory leading to very high ASA levels, while the second dose resulted in a decline in the ASSA levels, probably due to anergy.

### Time gap of 10-14 weeks for the second vaccine dose is beneficial

During the study period, the Indian government guidelines for the second vaccination dosage evolved from 4 weeks to 6-8 weeks and subsequently to 12-14 weeks. So we analysed the ASSA levels among the different time points in our CN group as their immune system had no memory to the SARS-CoV2 proteins. A small subset of seven HCWs in the CN group had taken their second vaccine dose between 10-14 weeks. For the analysis, we selected seven HCWs who were age-matched and ASSA level-matched on the 14^th^ day post-first vaccine dose, and had taken their second dose on 4 weeks or 6-8 weeks each. The GM ASSA levels were 5 U/ml for all the three groups on the 14^th^ day post-first vaccine dose. The rise in ASSA levels on the 14^th^ day after the second vaccine dose was analysed (Figure 3). The ASSA levels were the highest when the second dose was taken 10-14 weeks after the first dose. This difference was statistically significant (p=0.02; one-way Anova test).

**Figure 3:**
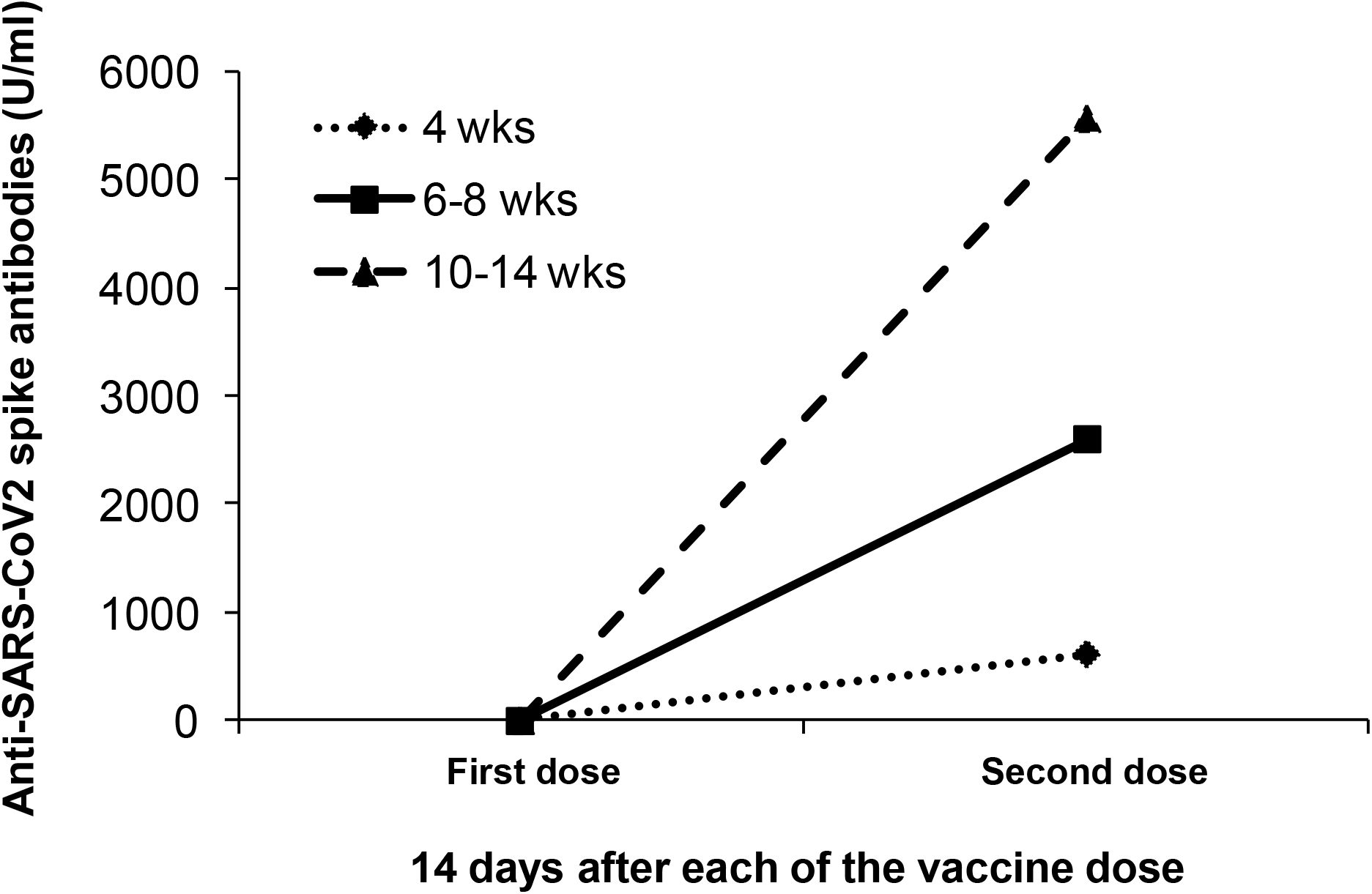
Longer time gap between the first and second vaccine doses is beneficial. The X-axis denotes the 14^th^ day sample after the first and second doses. The Y-axis denotes the anti-SARS-CoV2 spike antibody concentration in U/ml. The diamond with dotted line represents a 4-week time gap. The square with a solid line represents a 6-8 weeks time gap. The triangle with dashed line represents a 10-14 weeks time gap.

## Discussion

Determinants used to evaluate vaccine-mediated protection include measurement of antigen-specific antibodies; quality of such antibodies such as avidity, specificity or neutralization capacity; long-term persistence of these antibodies above the protective thresholds; and maintenance of immune memory cells that can respond rapidly in case of subsequent microbial exposure. In the current study, we determined the ASSA levels from the 14^th^ day post-first vaccine dose to three months post-second vaccine dose. We have shown that the ASSA levels peak on the 14^th^ day post-second vaccine dose and decline within three months thereafter in the CN group. Overall, the responder rate to Covishield was 83% on day 14, 98% on day 28 after the first dose, and 100% on day 14 after the second dose. Our results were similar to other studies after the first dose of ChAdOx1 [8,9].

In our study, the ASSA responses in the CN HCWs (GM – 1206 U/ml) at 14 days after the second dose were 6-fold higher than the PCPs (GM – 209 U/ml). The ASSA response after a natural infection is much less than that shown in other studies [10]. After the second dose, there was a 27-fold increase in the ASSA levels on the 14^th^ day in the CN group. Thus the first dose of Covishield elicited an antibody response as good as that of the natural infection among the South Indian population. Our findings also clearly indicate that the ASSA levels declined by 4-folds within three months after the second dose. The increase after the second dose was significantly greater when the second vaccine dose was taken 10-14 weeks after the first dose rather than four weeks. Our data support the current national recommendation for the time gap between the first and second doses. The longer time gap allows the activated cells to come back to the resting stage. A second dose at this time will work as a booster. On the other hand, if the second dose is given when the ASSA levels are high, it would result in anergy leading to negligible or small increase in antibody levels.

In the previously SARS-CoV2 infected mild (MCP) and asymptomatic (PEP) groups, the first vaccine dose boosted very high responses on the 14^th^ day. These responses only declined steadily up to the three months after the second dose. Throughout the longitudinal study period, the ASSA levels were 1.2 to 1.5-fold higher in the MCP group (mild) compared to the PEP group (asymptomatic). This indicates that the boosted ASSA responses were higher when the previous disease severity was higher. Similar to our findings, Velasco *et al* showed that the booster effect of the second dose was inversely proportional to the disease severity prior to the vaccination, and the booster effect was lower in the groups with a stronger response to the first prime dose [10]. These findings together suggest that in the previously infected groups the first vaccine dose worked as a robust booster of the memory cells and the second dose probably resulted in anergy or activation induced cell death due to the continuous challenge of the same immunogen within a short period of time. On the 14^th^ day after the second dose even though the ASSA levels declined they were still 8-fold and 11-fold higher in the PEP and MCP groups compared to the CN group. This probably suggests that a single vaccine dose works as a sufficient booster in the previously infected asymptomatic or mild groups. Additionally, checking the ASSA levels prior to the second dose might be beneficial to decide on the time gap between the two doses.

Additionally, in our study, we had 14% of BTI, most of which were after 28 days of the second vaccine dose when the GM of the ASSA levels was 1007 U/ml. This ASSA GM was almost 5-fold higher than the convalescent levels in the PCP group. The GM of ASSA levels at three months increased by 15-folds in the BTI group. However, all the HCWs had only mild symptoms and were managed as out-patients. Few other studies have shown similar BTI that are asymptomatic or mildly symptomatic after ChAd0X1 and the BNT1262b2, in spite of high antibody responses [1,10,11]. This suggests that the presence of ASSA at the time of BTI reduced the clinical severity of the disease.

In conclusion, our study delineates the longitudinal trend of the ASSA responses in vaccinees over a period of three months post-second dose both in CN and previously exposed HCWs. There is a clear benefit in giving the booster dose at least 3-4 months after the first priming dose. In the previously infected population, we have clearly shown that the first dose already acted as a booster. Since a number of HCWs could have been exposed asymptomatically to SARS-CoV2, it might be beneficial to check the ASSA levels before deciding on the time of the second dose. The ASSA levels declined considerably within three months suggesting that the ASSA are short-lived. However, future studies are warranted to determine further decline in later time points like six and 12 months that are crucial to understand the level of secondary and memory B cell responses to this vaccine. Additionally, it is also important to understand whether these ASSA would be present locally in the salivary and nasal glands to provide immediate protection against the invading SARS-CoV2.

## Data Availability

Not applicable

## Acknowledgements

Not applicable

## Availability of data and material

Not applicable

## Code availability

Not applicable

## Ethics approval

Yes. The study was approved by the VHS-Institutional Ethics Committee. The proposal number is VHS-IEC/72-2020.

## Consent to participate

Yes. Written informed consent has been obtained from all the participants.

## Consent to publish

Not applicable

